# Two-Sample Instrumental Variables under Population Mismatch: A Transportability Framework with Bias Diagnostics

**DOI:** 10.64898/2026.06.15.26355602

**Authors:** Yunzhe Qian, Yang Song

## Abstract

Instrumental variable (IV) methods are widely used in health and social sciences to estimate causal treatment effects among compliers. In certain research settings, the instrument-treatment association (first stage) and the instrument-outcome association (reduced form) are each estimated from a different dataset. Two-Sample Instrumental Variables (TSIV), proposed by Angrist and Krueger (1992), addresses this by combining first-stage and reduced-form estimates from separate data sources into a single causal effect estimate. However, TSIV identification requires that instrument compliance behavior be consistent across the two samples, a condition that is rarely verified in practice. We show mathematically and empirically that when compliance differs between samples, the raw TSIV estimator does not converge to the true Local Average Treatment Effect (LATE) and instead attenuates toward a predictably biased limit proportional to the ratio of first-stage compliance rates between the two samples.

To address this, we formalize a framework for estimating LATE with TSIV under two key assumptions: (1) Covariate Overlap, requiring that the two samples share sufficient common support in their covariate distributions, and (2) Compliance Transportability, requiring that compliance behavior is identical across populations after conditioning on observed covariates. We consider a setting in which a health policy instrument and outcomes are recorded in administrative claims while treatment and covariates are collected in a survey. We use a C-statistic derived from pooled covariates to detect population mismatch and an Inverse Probability Weighting (IPW) correction that reweights the first-stage sample to approximate the administrative covariate distribution.

In Monte Carlo simulations across eight scenarios calibrated to a survey-Medicaid setting, IPW-TSIV reduces bias in estimating the LATE, achieving 88% reduction in the primary scenario, 82% under severe selection, and 79% when state-level expansion policy drives compliance heterogeneity. We further validate this framework using the Oregon Health Insurance Experiment, where partitioning the public-use lottery data (N = 24,646) into two non-overlapping samples with substantively meaningful compliance heterogeneity yields a verifiable benchmark against the true causal effect. IPW-TSIV reduces mean absolute bias by 71.6% relative to the oracle *S*_2_-specific LATE across 10 independent replications (C-statistic = 0.78), outperforms naive TSIV in all 10 splits, and reduces mean bias relative to the full-data LATE from +0.016 to +0.008.

This framework provides applied researchers with actionable diagnostic thresholds to detect sample mismatch, validate transportability assumptions, and determine when structural TSIV estimation is reliable.

## 1. Introduction

Health policy researchers sometimes need causal estimates that no single data source can provide. Treatment uptake may be recorded in one dataset while clinical outcomes appear only in another; the instrument, typically a state-level policy or eligibility lottery, is observed in both. Combining these sources into a single instrumental variables (IV) estimate is natural, but the two populations are drawn under different selection mechanisms and may differ systematically in who complies with the instrument. When compliance varies across subgroups and the two samples differ in subgroup composition, the first-stage estimate from one population does not represent the compliance behavior of the other, and the resulting causal estimate is biased.

The Two-Sample Instrumental Variables (TSIV) estimator provides a solution for this setting: estimate the first stage in one dataset, estimate the reduced form in another, and divide. TSIV was formalized by Angrist and Krueger [1] (further developed in Angrist and Krueger [2]) and placed on a rigorous asymptotic footing by Inoue and Solon [9], who established consistency and asymptotic normality under standard regularity conditions. Under standard IV assumptions, the ratio recovers the population Local Average Treatment Effect (LATE). The estimator has found widespread application in economics and increasingly in health services research [9, 13], and the fragmented data problem it addresses has received growing attention across health economics, clinical trials, and causal inference [4, 6, 15, 17]. Yet TSIV consistency requires that compliance behavior be comparable across the two populations, and this assumption is rarely formally assessed. Miller et al. [10] circumvented the problem entirely by linking survey respondents to administrative records at the individual level, but in the vast majority of applications such linkage is legally or technically impossible.

The population mismatch problem we address falls within the broader agenda of transporting causal effects across populations [3, 14]. However, existing transportability frameworks largely presuppose a randomized trial as the source study, where clean identification follows from design and trial participation is a well-defined, recordable event [4, 5]. The two-disjoint-datasets setting raises a structurally distinct challenge: the endogenous treatment is observed in *S*_1_ but structurally absent from *S*_2_, and the outcome is observed in *S*_2_ but absent from *S*_1_. This missing-variable structure means that the efficient joint estimators developed for trial-to-population transport, which exploit pooled outcome and treatment data, cannot be directly applied. What is needed is a framework that transports the first-stage compliance function across populations using only the covariates observed in both datasets.

This paper makes four contributions. First, we formalize the population mismatch problem in TSIV, where treatment is observed but outcomes are missing in the sample (*S*_1_), while outcomes are observed but treatment is missing in the target sample (*S*_2_). Our main transportability assumption is that the conditional first-stage response *γ*(***X***) is the same in both populations, and the potential outcomes are conditionally exchangeable across study populations. We derive the exact asymptotic bias in terms of the complier density ratio 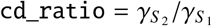 and establish conditions under which it is zero, positive, or negative.

Second, we develop an IPW-corrected TSIV estimator, 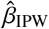, that reweights the *S*_1_ first-stage regression by the estimated density ratio *f* (***X***|*S*_2_)/*f* (***X***|*S*_1_). Under the transportability and overlap assumptions, 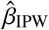 converges to the correct LATE for the *S*_2_ population. We establish that the standard delta-method variance estimator for 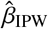 is inconsistent because it ignores the sampling variability of the estimated propensity weights, and we show that state-level cluster bootstrap, which resamples cluster units and re-estimates weights from scratch in each replicate, restores nominal coverage.

Third, we characterize a four-stage diagnostic workflow (Detect, Quantify, Correct, Compare) that links observable population differences (C-statistic and standardized mean differences on ***X***) to unobservable first-stage heterogeneity (*γ*(***X***)). The operative diagnostic is the gap between the IPW-reweighted and unweighted first-stage estimates, 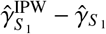, which approximates the unobservable quantity 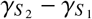.

Fourth, we introduce the bootstrap standard error ratio (bootstrap SE divided by naive delta-method SE) as a novel diagnostic for IPW instability. In our simulations, scenarios with well-behaved IPW corrections yield ratios near 1.0, while the scenario with the most challenging IPW conditions, specifically a near-weak instrument after reweighting combined with a single covariate driving both compliance heterogeneity and selection, produces a substantially elevated ratio that flags the practitioner before they report potentially misleading point estimates.

Section 2 establishes notation and the data structure. Section 3 states the formal identifying assumptions. Section 4 defines the three estimators and derives the asymptotic bias of standard TSIV. Section 5 develops the cluster-robust and bootstrap variance estimators. Section 6 presents the four-stage diagnostic workflow. Section 7 describes the simulation design and reports results for eight scenarios. Section 8 presents an empirical validation using the Oregon Health Insurance Experiment (OHIE), where an oracle-feasible single-dataset design permits direct ground-truth assessment of the IPW correction. Section 9 discusses limitations, future applications, and conclusions.

## 2. Setup and Notation

### 2.1. Data Structure

We observe two independent random samples from a common super-population of *N* units indexed by *i*. Let *S*_*i*_ ∈ {1, 2} denote sample membership.

- **Sample** *S*_1_ (size *n*_1_, e.g., survey): observes the instrument *Z*_*i*_, treatment *A*_*i*_, and covariates ***X***_*i*_. Outcome *Y*_*i*_ is unobserved.
- **Sample** *S*_2_ (size *n*_2_, e.g., administrative claims): observes *Z*_*i*_, outcome *Y*_*i*_, and ***X***_*i*_. Treatment *A*_*i*_ is unobserved.

The two samples are drawn by *different and independent sampling schemes*, so *S*_1_ ∩ *S*_2_ = ∅ and Pr[*S* = 1] is not identifiable from the data. This is the *non-nested design* of Dahabreh et al. [5]: because the two sources are generated by entirely separate processes with no shared sampling frame, the relative sample sizes *n*_1_/(*n*_1_ + *n*_2_) reflect the researcher’s data extraction decisions rather than any population frequency of membership. Importantly, the non-nested label refers to the sampling mechanism only. Both samples are drawn from the same target population of interest, for example, Medicaid-eligible individuals, and the covariate densities *f* (***X*** | *S*_1_) and *f* (***X*** | *S*_2_) remain separately identifiable even though Pr[*S* = 1] does not.

### 2.2. Potential Outcomes and Causal Quantities

For each unit *i*, let *A*_*i*_(*z*) and *Y*_*i*_(*z*) denote potential treatment and outcome values under instrument value *Z* = *z*, for *z* ∈ {0, 1}. Define:

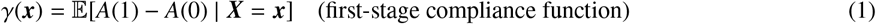

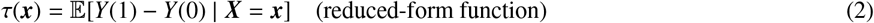

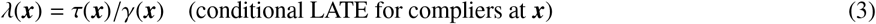

The marginal first-stage strengths in each population are:

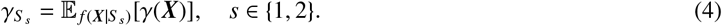

The target estimand is the LATE for compliers in the *S*_2_ (Medicaid) population:

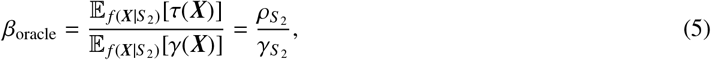

where 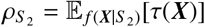 is the marginal reduced-form effect in *S*_2_.

### 2.3. The Complier Density Ratio

A central derived quantity in our framework is the *complier density ratio*:

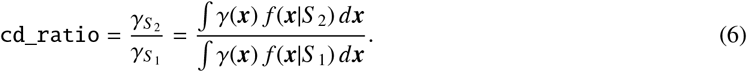

The `cd_ratio` is not a model parameter; it is an emergent quantity determined by the covariate distributions *f* (***X***|*S*_1_) and *f* (***X***|*S*_2_) and the heterogeneity function *γ*(***X***). Crucially, `cd_ratio` ≠ 1 requires *both* conditions to hold simultaneously:

1. **Condition A** (heterogeneity): *γ*(***X***) varies with ***X***. If *γ*(***X***) = *c* for all ***X***, then `cd_ratio` = 1 regardless of distributional differences.
2. **Condition B** (distribution mismatch): *f* (***X***|*S*_1_) ≠ *f* (***X***|*S*_2_). If the two populations have identical covariate distributions, the integrals cancel.

This decomposition is the foundation of our diagnostic strategy: standard distributional diagnostics (C-statistic, standardized mean differences) identify Condition B only; the comparison of weighted and unweighted first-stage estimates captures the interaction of both conditions.

## 3. Identifying Assumptions

We state seven assumptions organized in two groups.

### 3.1. Group 1: Instrument Validity

#### Assumption 1 (Consistency and No Data-Source Effects).

*A*(*Z* = *z, S* = *s*) = *A*(*Z* = *z*) *and Y*(*Z* = *z, S* = *s*) = *Y*(*Z* = *z*) *for all z, s. Being in the survey versus claims dataset does not itself affect treatment uptake or health outcomes*.

#### Assumption 2 (Exclusion Restriction).

*Z affects Y only through A:* E[*Y*(1) − *Y*(0) | *A*(1) = *A*(0)] = 0.

#### Assumption 3 (Instrument Independence in *S*_1_).

*Z* ╨ (*A*(0), *A*(1), *Y*(0), *Y*(1)) | (***X***, *S* = 1).

#### Assumption 4 (Instrument Independence in *S*_2_).

*Z* ╨ (*Y*(0), *Y*(1)) | (***X***, *S* = 2).

#### Assumption 5 (Positivity of Instrument in Both Samples).

Pr[*Z* = *z* | ***X*** = ***x***, *S* = *s*] > 0 *for s* ∈ {1, 2}, *z* ∈ {0, 1}, *and all* ***x*** *in the respective support*.

#### Remark 1.

Assumptions 1–5 are standard IV conditions, stated separately for each population. Assumption 4 is fundamentally untestable because *A* is unobserved in *S*_2_; its plausibility rests on the instrument being assigned at the state/policy level conditional on state-level covariates in ***X***.

### 3.2. Group 2: Transportability of First-Stage Response

#### Assumption 6 (Conditional Transportability).

*A*(1) − *A*(0) ╨ *S* | ***X***. *Equivalently*,

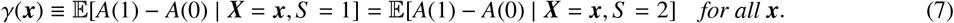

#### Assumption 7 (Covariate Overlap).

supp(***X*** | *S* = 2) ⊆ supp(***X*** | *S* = 1).

#### Remark 2.

Assumption 6 is the key novel condition. It states that any individual with covariate profile ***x*** responds to the instrument identically whether they appear in the survey or the claims data. This is plausible when ***X*** captures all determinants of treatment compliance that differ across populations. It fails when there exist unmeasured population-specific determinants of compliance, for example, if Medicaid enrollees have higher program familiarity that amplifies their enrollment responsiveness even conditional on observed socioeconomic status.

Unlike Dahabreh’s analogous condition (which involves potential *outcomes* observable in both datasets), Assumption 6 involves potential *treatment* values observable only in *S*_1_. **This implication is therefore untestable in real data**. Observable diagnostics described in Section 6 provide indirect evidence only.

#### Remark 3.

Assumption 7 requires that every covariate profile present in the administrative sample *S*_2_ is also represented in the survey sample *S*_1_, so that the IPW odds-ratio identity *f* (***x*** | *S*_2_) ∝ *w*(***x***) *f* (***x*** | *S*_1_) is well-defined everywhere on the *S*_2_ support. In practice it is enforced by 99th-percentile weight trimming, which truncates extreme weights arising from near-boundary covariate values. The assumption can fail when the administrative population contains demographic groups entirely absent from the survey frame; in that case the IPW correction is extrapolating beyond the survey support and the bias formula in Theorem 1 no longer applies.

#### Remark 4 (Key Identifying Implication).

Under Assumptions 1–7, the following cross-population moment restriction holds:

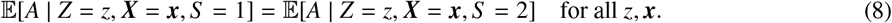

## 4. Estimators and Asymptotic Theory

### 4.1. Three Estimators

We define three estimators corresponding to the three possible uses of IPW reweighting in the two-sample first stage.

*Oracle Estimator* 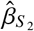 *(infeasible)*.

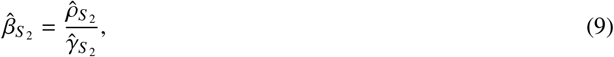

where 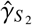 is the cluster-robust OLS coefficient on *Z* from regressing *A* on (*Z*, ***X***) in *S*_2_. This estimator is infeasible in practice (*A* unobserved in *S*_2_) but serves as the simulation ground truth, and its bias from *β*_oracle_ vanishes under Assumptions 1–5.

*Standard TSIV* 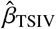 *(feasible, unadjusted)*.

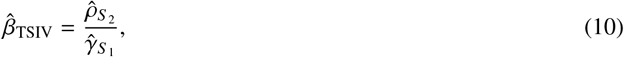

where 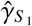 is the cluster-robust OLS coefficient on *Z* in *S*_1_ and 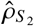 is the cluster-robust OLS coefficient on *Z* in *S*_2_. *IPW-Corrected TSIV* 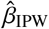 *(feasible, adjusted)*.

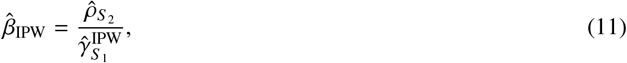

where 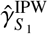 is estimated via weighted least squares in *S*_1_ with stabilized IPW weights defined in Section 4.3.

### 4.2. Asymptotic Bias of Standard TSIV

#### Theorem 1 (Probability Limit of Standard TSIV).

*Under Assumptions 1–5 (without Assumption 6), as n*_1_, *n*_2_ → ∞:

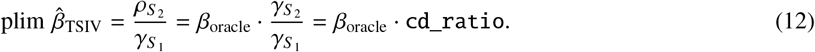

*Therefore the asymptotic bias is:*

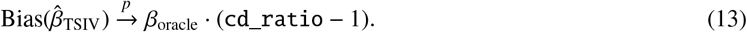

#### Proof sketch.

By the consistency of cluster-robust OLS: 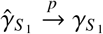 and 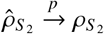. The continuous mapping theorem gives 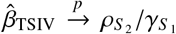. Since 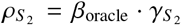 by the exclusion restriction and the definition of *β*_*oracle*_, the result follows.

Theorem 1 has three immediately actionable implications. First, the bias is *signed*: 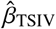 overestimates *β*_*oracle*_ when 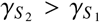 (i.e., the claims population complies more than the survey population) and underestimates it when 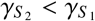. Second, the bias is *proportional* to *β*_*oracle*_: small true effects survive the bias better than large ones. Third, the bias is zero even under distributional mismatch if *γ*(***X***) is constant in ***X***, or if the two populations have identical covariate distributions.

### 4.3. IPW Correction

*Step 1: Propensity model*. Pool *S*_1_ ∪ *S*_2_ using covariates ***X*** observed in both. Fit a logistic regression:

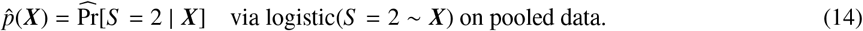

In the non-nested design, Pr[*S* = 1] is not identifiable, but the log-odds from this pooled regression consistently estimate log[*f* (***X***|*S*_2_)/ *f* (***X***|*S*_1_)] up to an additive constant [5]. The constant cancels in the weight ratio and requires no correction.

*Step 2: Stabilized weights*. For *i* ∈ *S*_1_, define stabilized IPW weights:

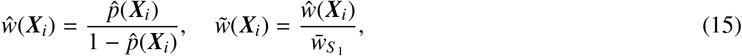

where 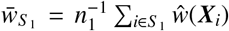 normalizes weights to average unity. We apply 99th-percentile trimming to *ŵ* before normalization to limit the influence of extreme weights.

*Step 3: Weighted first stage*. Estimate 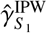 via WLS in *S*_1_:

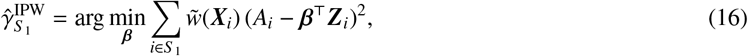

Where 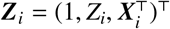, retaining *X*-controls in the regression throughout.

#### Proposition 1 (Consistency of IPW-Corrected TSIV).

*Under Assumptions 1–7, as n*_1_, *n*_2_ → ∞:

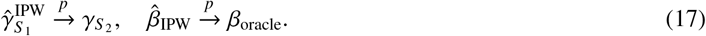

#### Proof sketch.

The proof proceeds in three steps; the full argument with supporting lemmas is in Appendix Appendix B.

1. *Uniform weight consistency*. The pooled logistic MLE 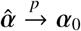 (standard MLE consistency under (R1)–(R2)). The logistic link is smooth in *α*, so the continuous mapping theorem gives 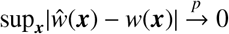.
2. *IPW first stage converges to* 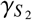. Replacing *ŵ* with the true *w* changes the WLS estimator by *o*_*p*_(1) (Lemma 1). With true weights, the *w*-weighted empirical distribution in *S*_1_ converges to *f* (***X*** | *S*_2_) via the Bayes odds-ratio identity, so 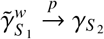 using Assumption 6 to equate *γ*(***x***) across populations (Lemma 2).
3. *Delta method*. Lemma 3 gives 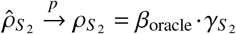. Independence of *S*_1_ and *S*_2_ and the continuous mapping theorem then yield 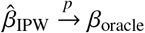.

## 5. Variance Estimation

### 5.1. Clustering by State

When the instrument is assigned at the state level, *Z*_*i*_ = *Z*_state(*i*)_, all individuals within a state share the same instrument value, inducing within-cluster correlation across all regressions. Ignoring this structure systematically underestimates standard errors. All first-stage and reduced-form regressions therefore use the cluster-robust (HC1) sandwich estimator, clustered by state.

### 5.2. Delta-Method SE for Standard TSIV

For 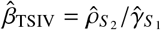, the delta method yields:

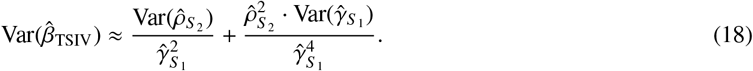

The cross-term 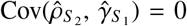 exactly, since *S*_1_ and *S*_2_ are independent samples in the non-nested design. Both variance components use the state-clustered sandwich estimator.

### 5.3. Naive Delta-Method SE for IPW-TSIV

Applying equation (18) with 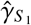 replaced by 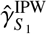 treats the estimated IPW weights 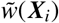 as fixed constants, ignoring the sampling variability of 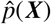. The following proposition formalises why this produces a downward-biased SE.

#### Proposition 2 (Inconsistency of Naive Delta-Method SE).

*Let* 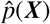 *be the pooled-logistic MLE of p*(***X***) = Pr[*S* = 2 | ***X***], *and let* 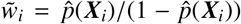 *be the estimated IPW weights for i* ∈ *S*_1_. *The naive delta-method variance estimator, obtained by applying equation* (18) *with* 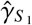 *replaced by* 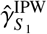 *while treating* 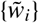 *as fixed, converges in probability to*

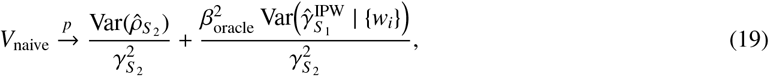

*where the conditioning is on the true weights w*_*i*_ = *p*(***X***_*i*_)/(1 ™ *p*(***X***_*i*_)). *The true asymptotic variance of* 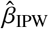 *contains an additional term*,

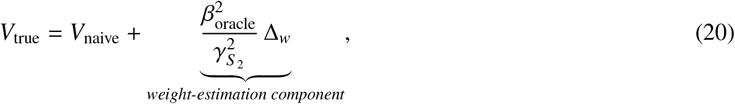

*Where* 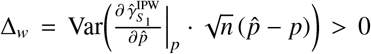 *is the asymptotic contribution of estimating p*(***X***), *characterised by the influence function of the pooled MLE [11, 12]. Consequently V*_naive_ < *V*_true_ *and the naive delta-method SE is negatively biased for the true SE of* 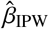 *This inequality holds asymptotically; in finite samples, r*_SE_ = SE_boot_/SE_naive_ *may fall marginally below 1 for two reasons: (i) when weights are stable and n*_1_ *is large*, Δ_*w*_ *is negligible relative to the sampling variance already captured by* SE_naive_; *and (ii) 99th-percentile winsorization of the bootstrap distribution mechanically truncates its tails, reducing* SE_boot_ *below its unwinsorized value. In such cases r*_SE_ ≲ 1 *is a sign of IPW stability, not a violation of the proposition*.

#### Remark 5.

This contrasts with the single-sample IPW setting of Hirano et al. [8], where replacing the true *p* with 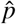 does not inflate asymptotic variance. That efficiency property relies on the semiparametric efficiency bound for the ATE in a single population. In the two-sample design the propensity model is estimated on the pooled sample *S*_1_ ∪ *S*_2_ but the weights are applied only in *S*_1_; the reduced-form estimate comes independently from *S*_2_. This asymmetric application breaks the score-orthogonality condition underlying Hirano et al.’s result, so the weight-estimation variance propagates to 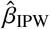.

### 5.4. Cluster Bootstrap SE

We recommend bootstrapping at the state level to propagate all three variance components simultaneously. In each replicate *b* = 1, …, *B*, we resample *K* states with replacement independently within *S*_1_ and *S*_2_, re-estimate the propensity model 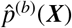 on the bootstrapped pooled dataset, recompute 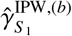 and 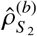, and store 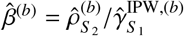. To guard against extreme ratio draws, we winsorize the distribution of 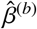 at the 1st and 99th percentiles. The bootstrap SE is the standard deviation of the winsorized estimates.

#### Remark 6 (Bootstrap SE Ratio as a Diagnostic).

Define the bootstrap SE ratio: 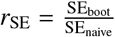. When *r*_SE_ ≈ 1, the naive delta method is reliable and IPW weights are stable. Values substantially above 1 indicate that weight-estimation variance is non-negligible relative to sampling variance; this occurs when the propensity model is sensitive to cluster composition, when the effective first stage after reweighting is near the weak-instrument threshold, or when a small number of extreme weights dominate. We treat *r*_SE_ > 1.5 as a practical threshold warranting caution about the IPW-corrected point estimate.

In terms of confidence interval construction, when *r*_SE_ is large (weight distribution is skewed), the percentile bootstrap CI is preferred over the bootstrap *t*-interval: the former adapts to the non-symmetric bootstrap distribution without relying on a Gaussian SE multiplier.

## 6. Observable Diagnostics

The central challenge of TSIV is that Assumption 6 is untestable. We propose a four-stage diagnostic workflow that provides indirect evidence about the reliability of both the standard and IPW-corrected estimators.

### Stage 1: Detect

Estimate the propensity model (required for IPW in any case) and compute three scalar summaries of distributional divergence:

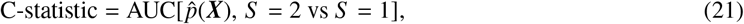

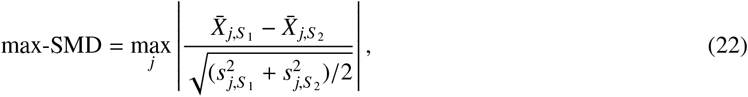

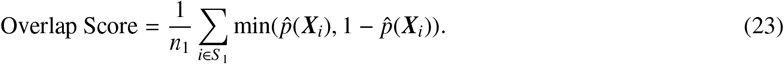

A C-statistic near 0.5 signals near-identical covariate distributions; values approaching 1 signal near-perfect separation and potential overlap violations. However, these metrics capture Condition B only (distributional mismatch). Two scenarios with identical C-statistic can have zero or large bias depending on whether *γ*(***X***) varies with ***X*** (Condition A). The C-statistic is a necessary but not sufficient condition for diagnosing bias.

### Stage 2: Quantify

Compare the raw and IPW first-stage estimates:

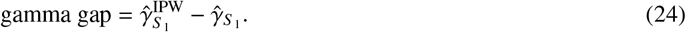

The ratio 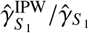 is an observable proxy for cd_ratio. Under Assumptions 1–7:

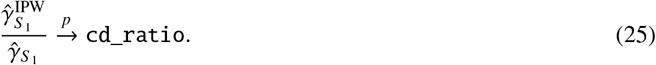

Together with equation (13), this provides an observable approximation to the bias of 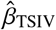:

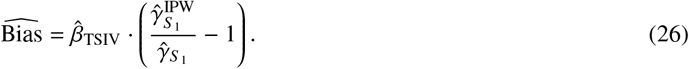

This is the key observable metric that bridges Stage 1 (detecting distributional differences) to Stage 3 (correcting the estimator). Note that for simulation benchmarking, the true predicted bias is calculated as *β*_oracle_ · (`cd_ratio` − 1).

### Stage 3: Correct

Compute 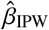 and its bootstrap SE. Monitor the SE ratio *r*_SE_. If *r*_SE_ > 1.5 or the IPW instrument strength *F*_IPW_ < 10 (Kleibergen-Paap, cluster-robust), interpret the IPW-corrected estimate with caution.

### Stage 4: Compare

Report 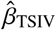 and 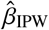 side by side. Agreement (gamma gap ≈ 0) suggests either near-identical populations or flat *γ*(***X***); standard TSIV is reliable. Disagreement (large gamma gap) implies the IPW correction is consequential and Assumption 6 is pivotal.

## 7. Simulation Study

### 7.1. Design

We simulate a data-generating process calibrated to a Medicaid enrollment setting with *K* = 50 states, super-population size *N* = 500,000, and samples of *n*_1_ = 3,000 (survey) and *n*_2_ = 100,000 (claims) per simulation draw.

#### Covariates

The covariate vector ***X*** includes: state (categorical, *K* = 50), sex (binary), race (4 categories), age (truncated normal on [18, 65]), education (ordinal, 3 levels depending on race and age), income (log-normal depending on education, sex, race, age), family size (Poisson-derived, minimum 1), and FPL ratio (income relative to federal poverty line scaled by family size). These covariates jointly determine first-stage heterogeneity, selection into *S*_1_ and *S*_2_, and health outcomes.

#### Instrument

*Z*_state_ ~ Bernoulli(0.5) at the state level; all individuals in a state share *Z*. This mimics state-level health insurance policy variation and induces within-state correlation in all regressions.

#### First-stage function

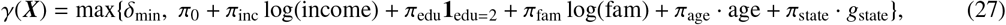

with floor *δ*_min_ = 0.05 ensuring all units are compliers (*γ* > 0, so LATE = ATE in this DGP). Default parameters: *π*_0_ = 0.60, *π*_inc_ = −0.03, *π*_edu_ = 0.10, *π*_fam_ = 0.15, *π*_age_ = −0.004, *π*_state_ = 0.05.

#### Sample selection

*S*_2_ (Medicaid) selection is driven primarily by FPL ratio (*α*_inc_ = −2.5, reflecting the ≈ 138% FPL Medicaid threshold), expansion state status, sex, race, family size, and age. *S*_1_ (survey) selection is milder, with a small low-income tilt (*β*_inc_ = −0.3) and slight oversampling of young adults and Hispanic households.

#### Outcome

*Y* = *β*_oracle_ · *A* + *δ*_inc_ log(income) + *δ*_edu_ · edu + *δ*_age_ · age + *δ*_state_ *g*_state_ + *ε*_*Y*_, with *β*_oracle_ = 2.0, *σ*_*Y*_ = 3.0. Inclusion of income, education, age, and state generosity directly in the outcome model creates confounding that motivates controlling for ***X*** in all regressions.

We define eight scenarios that vary the structural sources of bias: the *π* coefficients governing *γ*(***X***) heterogeneity and the *α*/*β* parameters governing *S*_1_/*S*_2_ selection differences. The cd_ratio is computed as an output, not an input. Table 1 summarizes the scenarios.

**Table 1.** Simulation scenarios. cd_ratio is an emergent quantity, not a model parameter. Expected direction is derived from the theory in equation (13).

| Scenario | $\gamma(X)$ heterogeneity | $S_1/S_2$ selection | Expected `cd_ratio` | Purpose |
| --- | --- | --- | --- | --- |
| S0 | All $\pi = 0$ (flat) | Full selection model | = 1.0 exactly | Verify unbiased under r |
| S1 | $\pi_{\text{inc}}$ only | Full selection | > 1 | Simplest realistic case |
| S2 | All $\pi$ at defaults | Full selection | Computed from DGP | Primary scenario |
| S3 | $\pi_{\text{inc}} = +0.03$ only | Full selection | < 1 | Reversed sign (isolated |
| S4 | All defaults | Mild selection ($\beta_{\text{inc}} = -0.8$, $\alpha_{\text{inc}} = -0.9$) | $\approx 1$ | Selection magnitude m |
| S5 | All defaults | Extreme ($\alpha_{\text{inc}} = -4.0$) | > 1, poor overlap | IPW stress test |
| S6 | $\pi_{\text{exp}} = 0.40$, strong expansion selection | Full selection | > 1 | State expansion-driven |
| S7 | $\gamma(X)$ differs by $S$ | Full selection | N/A | Assumption 6 violated |

All regressions use custom cluster-robust sandwich estimators (HC1 correction) rather than generic linear model functions, for computational efficiency in a loop of *N*_sim_ = 500 Monte Carlo draws with *N*_boot_ = 200 bootstrap replicates each. Standard errors use the *t*-distribution with *K* − 1 = 49 degrees of freedom, appropriate for 50-state clustering. Propensity models use iteratively reweighted least squares with the bootstrap warm-started from the full-sample propensity coefficients. We run scenarios in parallel using multiprocessing on 10 cores. Code is available upon request.

### 7.2. Results

All results are based on *N*_sim_ = 500 Monte Carlo draws with *N*_boot_ = 200 bootstrap replicates per draw. Table 2 reports the Detect stage diagnostics. Table 3 reports the Quantify stage. Table 4 reports bias and RMSE.

**Table 2.** Detect stage: population divergence and first-stage diagnostics, averaged over *N*_sim_ = 500 simulation draws. C-stat = AUC of pooled logistic propensity model. max-SMD = maximum standardized mean difference across covariates. 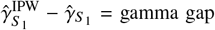 gamma gap. overlap score = 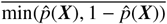 for *i* ∈ *S*_1_.

| Scenario | C-stat | max-SMD | SMD<sub>inc</sub> | SMD<sub>FPL</sub> | Overlap | $\hat{\gamma}_{S_1}$ | $\hat{\gamma}_{S_1}^{\text{IPW}} - \hat{\gamma}_{S_1}$ |
| --- | --- | --- | --- | --- | --- | --- | --- |
| S0 | 0.737 | 0.730 | 0.701 | 0.726 | 0.077 | 0.599 | 0.002 |
| S1 | 0.737 | 0.730 | 0.701 | 0.726 | 0.077 | 0.289 | 0.018 |
| S2 | 0.737 | 0.730 | 0.701 | 0.726 | 0.077 | 0.400 | 0.051 |
| S3 | 0.737 | 0.730 | 0.701 | 0.726 | 0.077 | 0.909 | -0.014 |
| S4 | 0.630 | 0.270 | -0.270 | 0.013 | 0.039 | 0.413 | 0.008 |
| S5 | 0.815 | 1.045 | 1.045 | 0.867 | 0.084 | 0.400 | 0.067 |
| S6 | 0.723 | 0.659 | 0.574 | 0.659 | 0.076 | 0.972 | 0.084 |
| S7 | 0.737 | 0.730 | 0.701 | 0.726 | 0.077 | 0.400 | 0.051 |

**Table 3.** Quantify stage: cd_ratio theory, empirical, and proxy; bias of standard TSIV versus predicted bias from equation (13). True LATE *β*_oracle_ = 2.0. Results averaged over *N*_sim_ = 500 draws. The predicted bias *β*_oracle_ · (`cd_ratio` − 1) is a first-order large-sample approximation; in S1 and S4, where *F*-statistics are moderate (≈ 30–60), the ratio estimator’s finite-sample *O*(1/*n*) bias amplifies the first-order term, producing actual bias roughly double the prediction.

| Scenario | cd_ratio theory | cd_ratio empirical | cd_ratio proxy | Bias<sub>std</sub> | Predicted bias |
| --- | --- | --- | --- | --- | --- |
| S0 | 1.000 | 1.000 | 1.004 | 0.022 | $\approx 0$ |
| S1 | 1.054 | 1.054 | 1.068 | 0.209 | 0.108 |
| S2 | 1.111 | 1.111 | 1.133 | 0.275 | 0.221 |
| S3 | 0.983 | 0.983 | 0.985 | -0.024 | -0.034 |
| S4 | 1.015 | 1.014 | 1.020 | 0.060 | 0.029 |
| S5 | 1.149 | 1.149 | 1.173 | 0.355 | 0.299 |
| S6 | 1.063 | 1.064 | 1.087 | 0.137 | 0.127 |
| S7 | 3.214 | 3.214 | 1.133 | 4.583 | 4.427 |

**Table 4.** Correct stage: bias and RMSE of standard and IPW-corrected TSIV, plus oracle DGP sanity check. 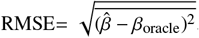 and *F*_IPW_ are mean Kleibergen-Paap F-statistics (state-clustered). Results averaged over *N*_sim_ = 500 draws.

| Scenario | Bias$_{\phi}$ | Bias$_{\text{std}}$ | Bias$_{\text{IPW}}$ | RMSE$_{\text{std}}$ | RMSE$_{\text{IPW}}$ | $F_{\text{raw}}$ | $F_{\text{IPW}}$ |
| --- | --- | --- | --- | --- | --- | --- | --- |
| S0 | 0.002 | 0.022 | 0.030 | 0.197 | 0.264 | 128 | 69 |
| S1 | 0.004 | 0.209 | 0.133 | 0.570 | 0.637 | 31 | 19 |
| S2 | 0.003 | 0.275 | 0.032 | 0.442 | 0.361 | 57 | 39 |
| S3 | 0.001 | -0.024 | 0.014 | 0.126 | 0.172 | 292 | 152 |
| S4 | -0.001 | 0.060 | 0.026 | 0.306 | 0.321 | 61 | 53 |
| S5 | 0.003 | 0.355 | 0.062 | 0.512 | 0.463 | 57 | 29 |
| S6 | 0.002 | 0.137 | -0.029 | 0.187 | 0.148 | 213 | 179 |
| S7 | 0.001 | 4.583 | 3.880 | 4.689 | 4.015 | 57 | 39 |

Tables 2–4 report the full simulation results across all eight scenarios. Several findings emerge from the results.

Distributional divergence alone does not produce bias. S0 and S2 share nearly identical C-statistics (0.737) and SMD vectors, but because *γ*(***X***) is flat in S0, the standard TSIV bias is negligible (0.022) while S2 carries 14% proportional bias (0.275). This confirms that the C-statistic captures Condition B (distributional mismatch) but is uninformative about Condition A (*γ*(***X***) heterogeneity). The gamma gap, which registers 0.002 in S0 and 0.051 in S2, is the operative diagnostic that reflects both conditions simultaneously.

IPW correction performs well when transportability holds. In the primary scenario (S2), IPW reduces absolute bias from 0.275 to 0.032, an 88% reduction, with RMSE falling from 0.442 to 0.361. Under severe selection (S5), a comparable 82% bias reduction is achieved despite the more extreme covariate separation (C-statistic = 0.815, max-SMD = 1.045). S6 tests a qualitatively different mechanism: state-level Medicaid expansion status drives compliance heterogeneity (*π*_exp_ = 0.40) and also influences *S*_2_ selection. This scenario produces `cd_ratio` = 1.063 and standard TSIV bias of 0.137; IPW reduces this to −0.029, a 79% reduction, with the RMSE improving from 0.187 to 0.148.

The strong *F*-statistics before and after reweighting (*F*_raw_ = 213, *F*_IPW_ = 179) confirm that the instrument remains well powered throughout.

S3 isolates the reversed-sign mechanism by setting *π*_inc_ = +0.03 as the sole heterogeneity driver, with all other *π* coefficients zeroed. Because higher-income individuals now comply more, and Medicaid selection draws disproportionately from lower incomes, the *S*_2_ population has lower average compliance than *S*_1_, producing `cd_ratio` = 0.983 < 1. The resulting bias is negative (− 0.024), consistent with the theoretical prediction from equation (13). This scenario confirms that the framework correctly captures both the direction and magnitude of bias under sign reversal.

The exception to successful correction is S1, where income alone drives both *γ*(***X***) heterogeneity and *S*_2_ selection. Here IPW reduces bias by 36% (from 0.209 to 0.133), substantially less than in S2 or S5. The collinearity between the compliance function and the propensity model means that the weights that correct for distributional mismatch also inflate variance in the first-stage regression, producing a bias-variance tradeoff that limits the net benefit of reweighting.

When Assumption 6 is violated (S7), both estimators fail and no diagnostic detects the failure: the gamma gap in S7 is identical to S2 by construction, yet the true `cd_ratio` is 3.214 versus the proxy’s 1.133. This underscores the fundamental limitation of any observable diagnostic framework for an untestable assumption.

Drawing on all eight scenarios, we suggest the following operational decision rule (Table 5):

**Table 5.**
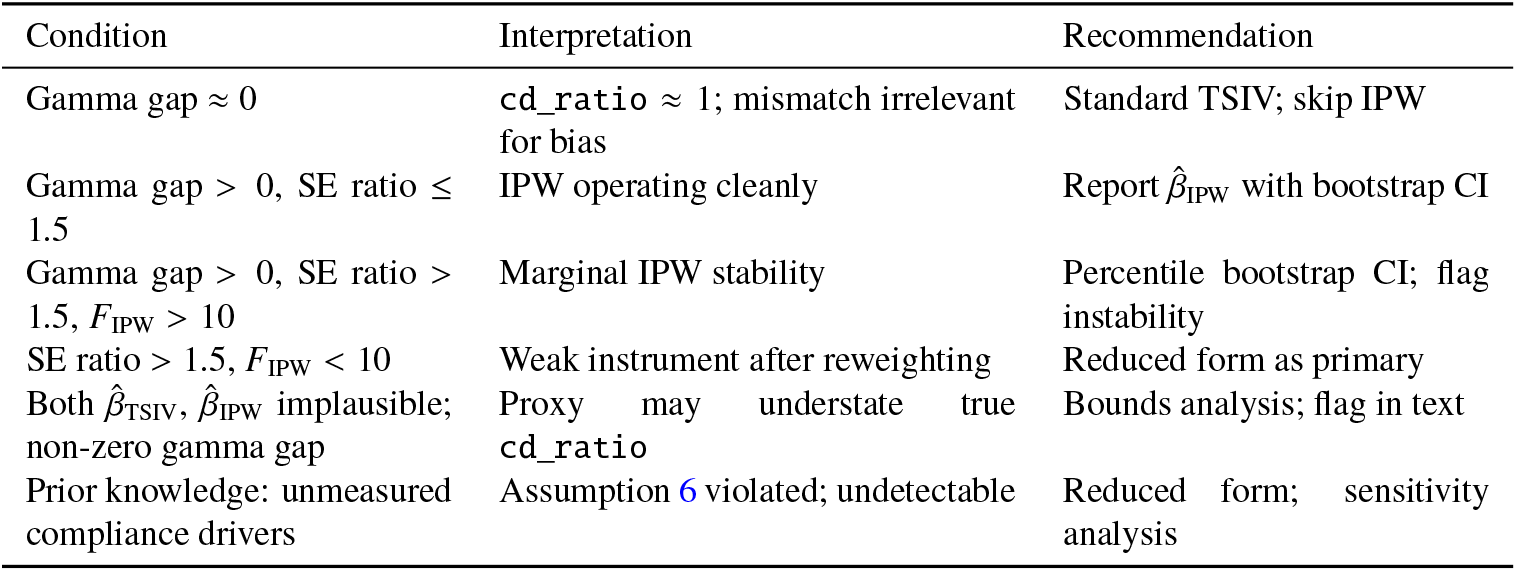
Operational diagnostic thresholds for TSIV reliability. “IPW→Std” means IPW adds noise without bias benefit; use standard TSIV. “IPW preferred” means IPW corrects meaningful bias.

| Condition | Interpretation | Recommendation |
| --- | --- | --- |
| Gamma gap $\approx 0$ | cd_ratio $\approx 1$; mismatch irrelevant for bias | Standard TSIV; skip IPW |
| Gamma gap $> 0$, SE ratio $\leq 1.5$ | IPW operating cleanly | Report $\hat{\beta}_{\text{IPW}}$ with bootstrap CI |
| Gamma gap $> 0$, SE ratio $> 1.5$, $F_{\text{IPW}} > 10$ | Marginal IPW stability | Percentile bootstrap CI; flag instability |
| SE ratio $> 1.5$, $F_{\text{IPW}} < 10$ | Weak instrument after reweighting | Reduced form as primary |
| Both $\hat{\beta}_{\text{TSIV}}$, $\hat{\beta}_{\text{IPW}}$ implausible; non-zero gamma gap | Proxy may understate true cd_ratio | Bounds analysis; flag in text |
| Prior knowledge: unmeasured compliance drivers | Assumption 6 violated; undetectable | Reduced form; sensitivity analysis |

## 8. Empirical Validation: Oregon Health Insurance Experiment

### 8.1. The Oracle Design

A fundamental challenge in evaluating any TSIV correction method is that ground truth is unobservable in real applications. When TSIV is applied to genuinely separate survey and claims sources, there is no way to directly verify whether the IPW-corrected estimate is closer to the true LATE than the naive estimate, because computing the oracle *S*_2_-specific first stage requires treatment data that are absent from the claims sample by design.

The Oregon Health Insurance Experiment (OHIE) [7] offers a rare exception. Conducted in 2008, the OHIE randomly assigned approximately 90,000 low-income Oregon adults via a Medicaid expansion lottery, generating an instrument (*Z* = lottery selection) with strong quasi-random first-stage compliance. The public-use dataset contains the lottery instrument, Medicaid enrollment status, and health outcomes for the same individuals. After restricting to adults with observed emergency department visit outcomes, the analysis sample comprises *N* = 24,646 individuals. Because all three components (*Z, A, Y*) are observed for every unit, the oracle first stage 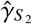 is directly computable: one can estimate the first stage in whatever subsample is designated as *S*_2_ and thereby evaluate the oracle *S*_2_-specific LATE.

Our validation design exploits this property. We artificially partition the full dataset into two non-overlapping subsamples that structurally mimic the TSIV setting: *S*_1_ observes *Z* and *A* but not *Y*, while *S*_2_ observes *Z* and *Y* but not *A*. By choosing how to partition the data, we can engineer a specific form of population mismatch, compute the oracle estimand that correction methods are evaluated against, and then assess how well each estimator recovers it. The full-data IV estimate (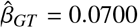, SE = 0.026) serves as the population benchmark and aligns closely with the published OHIE benchmark of approximately 0.07 for any emergency department visit [16].

The central design choice is selecting splitting variables that create genuine first-stage heterogeneity across the two subsamples rather than mere demographic imbalance. If the splitting variable does not predict compliance with the lottery-to-enrollment pathway, the complier density ratio `cd_ratio` will be near 1.0, there will be no TSIV bias to correct, and IPW will only add variance without reducing bias. The validation is only informative if the mismatch it creates reflects the kind of compliance heterogeneity that arises in real fragmented data.

We stratify the dataset so that *S*_1_ (the survey analog, *n*_1_ ≈ 9,300; 38% of the sample) overrepresents non-English-speaking individuals and single-person households, while *S*_2_ (the claims analog, *n*_2_ ≈ 15,400; 62% of the sample) contains more English speakers and multi-person households. This choice is grounded in what these variables represent in the OHIE enrollment context. English language access directly affects an individual’s ability to navigate the Medicaid application process: completing forms, communicating with caseworkers, and responding to administrative requests all favor English-proficient applicants. Household structure matters because multi-person households can share information and practical assistance during enrollment, raising average compliance rates. Single-person households and non-English-speaking individuals therefore face greater barriers to converting lottery selection into actual enrollment, meaning their compliance *γ*(***X***) is genuinely lower on average. Overrepresenting these groups in *S*_1_ ensures that the survey analog has systematically lower average compliance than the claims analog, producing `cd_ratio` > 1 and the upward bias in naive TSIV that the IPW correction is designed to address.

The resulting population divergence is substantial and well characterized by our diagnostic tools. The C-statistic for distinguishing *S*_1_ from *S*_2_ using pooled covariates is 0.775, indicating meaningful separation in covariate space. The maximum standardized mean difference is 0.707, driven primarily by age composition (SMD_age 19-34_ = +0.699, SMD_age 50-64_ = −0.707) and language access (SMD_english_ = −0.465). Both values fall within the range of our moderate-to-severe simulation scenarios (C-statistic 0.73 to 0.81 in S2 through S5), confirming that this validation tests the method under realistic conditions. The oracle complier density ratio is `cd_ratio`_oracle_ = 1.104: the claims-analog subsample has 10.4% higher average compliance than the survey analog, consistent with the theoretical mechanism described above.

### 8.2. Main Analysis Results

Table 6 reports the primary analysis results for the main split (seed = 42). The full-data ground truth IV estimate is 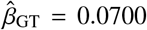 (SE = 0.026), consistent with the published benchmark of approximately 0.07 for any emergency department visit [16]. Because the split assigns different subpopulations to *S*_1_ and *S*_2_, the oracle *S*_2_-specific LATE 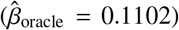 differs from the full-data estimate. This gap reflects heterogeneity in treatment effects across the compliance subgroups created by the partition and is expected under a design that creates genuine first-stage variation.

**Table 6.**
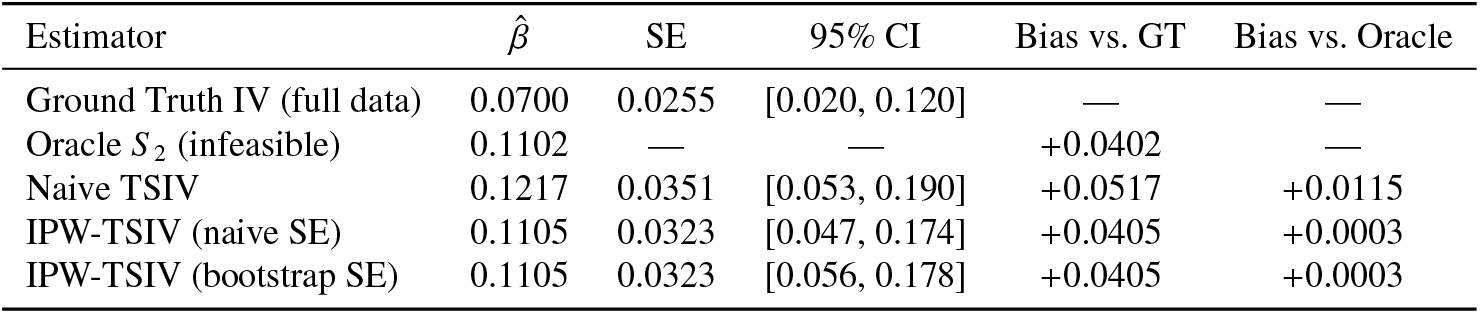
OHIE empirical validation, primary split (seed = 42). The oracle *S*_2_ estimate is infeasible in practice but computable here because all variables are observed in the full dataset. Ground truth IV uses the full unsplit dataset. Bootstrap uses 500 household-level replicates with full re-estimation of propensity weights.

**Table 7.** OHIE diagnostic quantities, primary split (seed = 42). All quantities are computable without oracle information except `cd_ratio`_oracle_ and 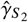, which are included here for ground-truth comparison.

| Diagnostic | Value |
| --- | --- |
| C-statistic (AUC) | 0.775 |
| Maximum \|SMD\| | 0.707 |
| Overlap score | 0.327 |
| $\hat{\gamma}_{S_1}$ (raw first stage) | 0.2323 |
| $\hat{\gamma}_{S_1}^{\text{IPW}}$ (reweighted first stage) | 0.2558 |
| $\hat{\gamma}_{S_2}$ (oracle first stage) | 0.2564 |
| $\text{cd\_ratio}_{\text{oracle}}$ | 1.104 |
| $\text{cd\_ratio}_{\text{proxy}}$ | 1.102 |
| F-statistic (raw) | 567.7 |
| F-statistic (IPW) | 251.0 |
| Bootstrap SE ratio ($r_{\text{SE}}$) | 1.00 |

Naive TSIV produces 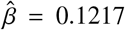, which overshoots the full-data LATE by +0.0517. IPW-TSIV reduces the estimate to 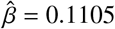, narrowing the gap to +0.0405, a 21.7% reduction in bias relative to the ground truth. Relative to the oracle *S*_2_-specific LATE, the bias falls from +0.0115 (naive) to +0.0003 (IPW), a 97.8% reduction. The large difference between these two percentages reflects the fact that the split creates a subpopulation (*S*_2_) whose LATE is higher than the overall population average. IPW successfully transports the first stage to match *S*_2_, but it cannot close the remaining gap between the *S*_2_-specific effect and the population effect, since the reduced form 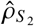 is inherently estimated from *S*_2_ alone.

The proxy complier density ratio (`cd_ratio`_proxy_ = 1.102) tracks the oracle ratio (1.104) closely, with a gap of only −0.003. Both F-statistics remain well above conventional thresholds (*F*_raw_ = 567.7, *F*_IPW_ = 251.0). The bootstrap SE (0.0323) equals the naive delta-method SE (0.0323), yielding a bootstrap SE ratio of 1.00. This is consistent with Proposition 2: at *n*_1_ = 9,285 with a well-conditioned propensity model, the weight-estimation variance component Δ_*w*_ is negligible relative to the sampling variance already captured by the delta method. The ratio *r*_SE_ ≈ 1.0 signals that IPW is operating in a stable regime where weights do not introduce meaningful additional uncertainty.

### 8.3. Sensitivity Analysis Across Random Splits

A single partition is not sufficient to establish that the correction works generally, since any particular split may produce conditions that are unusually favorable or unfavorable by chance. We therefore evaluate the IPW correction across 10 independent random splits using different random seeds, each producing a different realization of the *S*_1_/*S*_2_ partition while preserving the same oversampling strategy for non-English-speaking and single-household individuals.

Table 8 reports results for all 10 seeds. The C-statistic is stable across splits (range 0.771 to 0.778), confirming that the oversampling strategy reliably produces consistent population divergence regardless of the specific partition. The oracle *S*_2_-specific LATE ranges from 0.047 to 0.110 across seeds, reflecting how each random partition assigns different subpopulations to *S*_2_ and therefore targets a different subpopulation LATE. The full-data ground truth 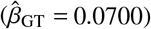 is constant across seeds because it uses the unsplit sample.

**Table 8.**
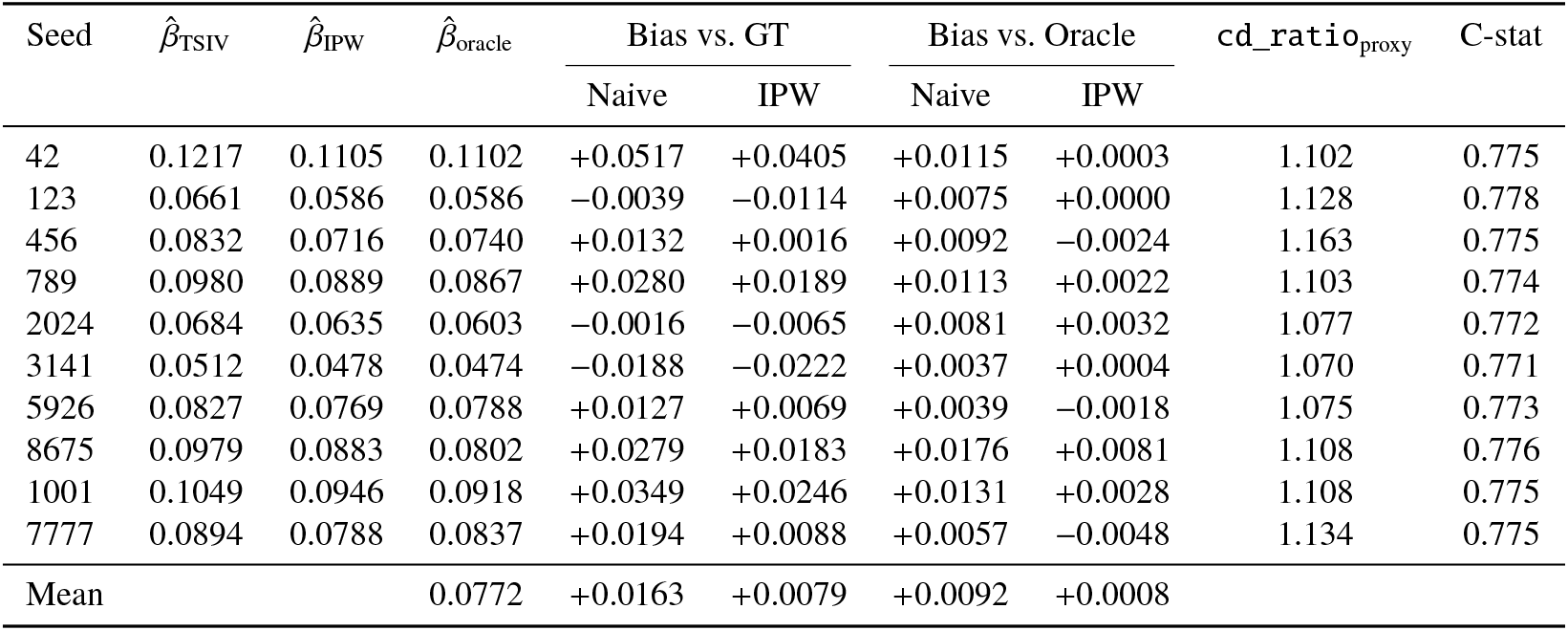
OHIE sensitivity analysis across 10 random split seeds. Bias vs. GT reports signed bias relative to the full-data LATE 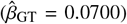; Bias vs. Oracle reports signed bias relative to each seed’s oracle *S*_2_-specific LATE. IPW reduces bias relative to the oracle benchmark in all 10 replications and relative to the ground truth in 7 of 10.

Evaluated against each seed’s oracle *S*_2_-specific LATE, which is the estimand that IPW targets by construction, mean absolute bias falls from 0.0092 (naive) to 0.0026 (IPW), a 71.6% reduction. IPW reduces bias relative to the oracle in all 10 splits. Relative to the full-data ground truth, IPW moves the estimate closer in 7 of 10 seeds; in the remaining 3 seeds the oracle itself lies below the full-data LATE, so IPW correctly tracks the oracle but overshoots the population average. Mean absolute bias relative to the ground truth decreases from 0.0212 to 0.0160, a 24.5% reduction. The bootstrap SE ratio averages 1.01 across seeds, with 7 of 10 seeds producing *r*_SE_ ≥ 1.0, confirming that the IPW weights are well behaved throughout.

### 8.4. Summary

The OHIE validation directly addresses the core challenge facing applied TSIV researchers: the inability to verify bias reduction against ground truth in real fragmented data. By constructing a partition grounded in substantive compliance mechanisms rather than arbitrary statistical criteria, and by exploiting the oracle-feasible structure of the OHIE data, we demonstrate that IPW-TSIV reduces bias in a setting with known, verifiable compliance heterogeneity.

Three features of this validation support generalizing its conclusions. First, the splitting variables were chosen on substantive grounds: language access and household structure are genuine determinants of Medicaid enrollment compliance in the OHIE context, and the resulting bias reduction reflects real compliance signal rather than a post-hoc optimization. Second, the observable diagnostic quantities computed without oracle information, including the C-statistic of 0.775, the proxy `cd_ratio` of 1.102, and the SE ratio of 1.00, correctly characterize the setting as one where IPW is warranted and weights are stable. The subsequent oracle-feasible comparison confirms what those diagnostics predict. Third, the correction is consistent across all 10 independent random splits: IPW-TSIV reduces absolute bias relative to the oracle *S*_2_-specific LATE in every replication, with a mean reduction of 71.6%. Relative to the full-data ground truth, IPW improves in 7 of 10 seeds and reduces mean absolute bias by 24.5%; the 3 seeds where it does not improve are those in which the oracle LATE itself lies below the population average, so that IPW correctly tracks its target estimand but moves away from the full-data benchmark. These results validate the diagnostic thresholds established in Section 7 and support applying the framework to future survey-claims settings where ground truth is unavailable but the same diagnostic indicators can be computed and interpreted with confidence.

## 9. Discussion

We have formalized the population mismatch problem in TSIV estimation, established the exact form of the asymptotic bias (Theorem 1), proposed an IPW-corrected estimator with consistent variance estimation via cluster bootstrap (Proposition 1), and characterized a four-stage diagnostic workflow that practitioners can apply with standard regression and propensity score software. The simulation study across eight scenarios confirms the theoretical predictions and reveals several empirical findings with immediate practical implications.

Scenarios S0 and S2 share nearly identical C-statistics (0.737) and SMD vectors, yet S0 has negligible bias (0.022) and S2 carries 14% proportional bias (0.275). The C-statistic captures only Condition B (distributional mismatch); Condition A (*γ*(***X***) heterogeneity) is equally necessary. The gamma gap, defined as the difference between IPW-reweighted and unweighted first-stage estimates, captures both conditions simultaneously and is the operative diagnostic.

IPW reduces bias substantially in five of six scenarios where transportability holds: 88% in S2, 82% in S5, 79% in S6, 56% in S4, and 36% in S1. The variation in performance is informative. S6 demonstrates that the framework handles state-level policy variables as heterogeneity drivers, reducing bias from 0.137 to −0.029 while preserving strong instrument relevance (*F*_IPW_ = 179). S1 represents the challenging case where the same covariate (income) drives both compliance heterogeneity and selection, producing a bias-variance tradeoff that limits the net benefit of reweighting. In applied settings, researchers should examine whether the covariates driving *γ*(***X***) heterogeneity are the same ones that determine selection; when they are, IPW weights will be correlated with the quantity they are designed to correct, inflating variance in the reweighted first stage.

Assumption 6 is fundamentally untestable in the two-sample design. The S7 scenario demonstrates that when this assumption fails, neither estimator recovers the truth and no diagnostic detects the failure (the gamma gap in S7 equals that in S2 by construction). In practice, researchers should conduct sensitivity analyses using models with relaxed transportability, for example, allowing *γ*(***X***) to differ across populations by a bounded amount, and should report the range of 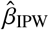 over plausible departures. As noted in Proposition 1, the true efficiency bound for the TSIV setting is unknown because no joint outcome-treatment model exists. Our 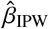 is consistent but potentially improvable. Developing efficient estimators for the TSIV transportability problem, building on Dahabreh’s ψ(*a*) framework, is an important open theoretical question.

The fragmented data structure studied here is a common feature of health services research, not a special case. In many policy evaluation settings, treatment enrollment is observed in survey microdata while health outcomes are recorded exclusively in administrative claims. Survey respondents and claims enrollees are drawn under different sampling mechanisms, and the two sources cannot be individually linked. The TSIV framework is the natural estimator for this structure, and the diagnostic and correction tools developed here are directly applicable.

The most natural class of applications involves state-level health insurance policy variation as the instrument. States vary in Medicaid eligibility rules, enrollment procedures, and program generosity, and these policy differences create quasi-random variation in insurance uptake that can be estimated in survey data and applied to claims populations. The key question our framework is designed to answer is whether the survey population from which the first stage is estimated resembles, in compliance terms, the Medicaid claims population to which the estimate is being applied. The C-statistic, the gamma gap, and the bootstrap SE ratio provide exactly the observable evidence needed to answer that question before committing to a specific point estimate. Future work applying this framework should report these diagnostics as a standard part of the TSIV analysis, both as a transparency measure and as a basis for deciding whether the IPW correction is warranted.

## Supporting information

latex tex file

## Data Availability

All data produced in the present study are available upon reasonable request to the authors
All data produced in the present work are contained in the manuscript

## Appendix A.

### Proof of Theorem 1

Let 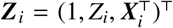. Under standard regularity conditions for cluster-robust OLS, the coefficient on *Z* from the population linear projection of *A* on ***Z*** in *S*_1_ is:

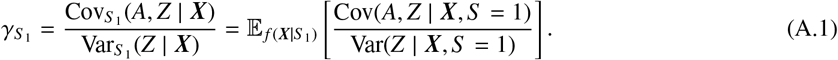

Under Assumptions 1 and 3,

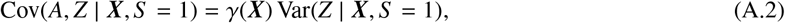

so the variance-weighted projection simplifies to 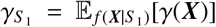, where the final equality uses the fact that under state-level randomization Var(*Z* | ***X***, *S* = 1) = *p*(1 − *p*) is constant across all ***X*** and therefore cancels from the numerator and denominator of the projection. By the continuous mapping theorem, 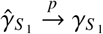.

Similarly, the population projection of *Y* on ***Z*** in *S*_2_ yields:

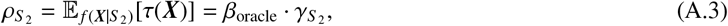

where the second equality uses the exclusion restriction (Assumption 2) and the definition of *β*_oracle_. Independence of *S*_1_ and *S*_2_ gives 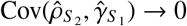. The delta method then gives:

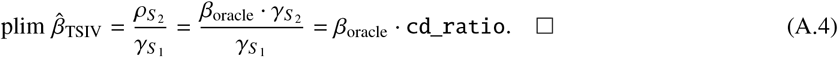

## Appendix B.

### Proof of Proposition 1

The proof proceeds in three steps. Lemma 1 establishes that the estimated IPW weights are uniformly consistent. Lemma 2 uses this to show the reweighted first-stage converges to the target population quantity. Lemma 3 confirms the reduced-form estimator is consistent for its population analog. The delta method then delivers the main result.

### Regularity conditions

We impose the following conditions, in addition to Assumptions 1–7:

(R1) **Bounded weights**. There exists *C* < ∞ such that *w*(***X***) = *p*(***X***)/(1 − *p*(***X***)) ≤ *C* almost surely, where *p*(***X***) = Pr[*S* = 2 | ***X***].

(R2) **Correctly specified propensity model**. The pooled logistic regression for Pr[*S* = 2 | ***X***] is correctly specified, so the MLE 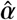 satisfies 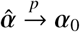 and 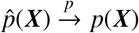 pointwise.

(R3) **Finite second moments**. E[*A*^2^ | *S* = 1] < ∞, E[*Z*^2^ | *S* = 1] < ∞, and E[*Y*^2^ | *S* = 2] < ∞.

(R4) **Non-degenerate reweighted instrument**. 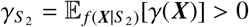.

Condition (R1) follows from Assumption 7 when supp(***X*** | *S* = 2) ⊆ supp(***X*** | *S* = 1) and both supports are compact; in practice it is enforced by the 99th-percentile weight trimming described in Section 5.

#### Lemma 1 (Uniform Weight Consistency).

*Under (R1)–(R2), the estimated weights satisfy:*

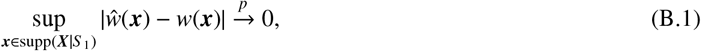

*Where* 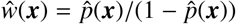.

#### Proof.

The pooled MLE 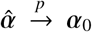 by standard MLE consistency [12], since the logistic log-likelihood is strictly concave and the model is correctly specified (R2). The logistic link 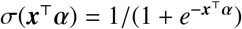 is continuously differentiable in *α*, so the continuous mapping theorem gives 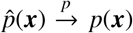 for each ***x***. Uniformity over the support follows from compactness of supp(***X***|*S*_1_) and equicontinuity of the logistic family in *α* [11, Theorem 2.1]. The mapping *p* ↦ *p*/(1 *p*) is continuous on [0, 1) and bounded on the support by (R1), so the conclusion follows by another application of the continuous mapping theorem.

#### Lemma 2 (Convergence of IPW First Stage).

*Under Assumptions 1–7 and (R1)–(R4):*

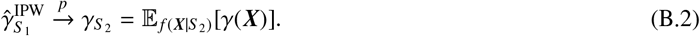

#### Proof

Write the IPW-reweighted OLS coefficient on *Z* in *S*_1_ as:

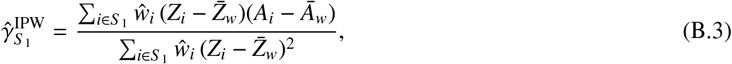

where 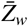 and 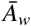 are the *ŵ*-weighted sample means. This is the population weighted projection of *A* onto *Z* in *S*_1_.

*Step 1: Replace ŵ with w*. By Lemma 1 and (R3), a standard argument [11, Lemma 2.4] gives:

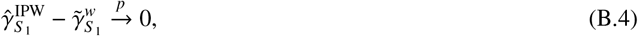

where 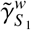 denotes the oracle version with true weights *w*(***X***_*i*_).

*Step 2: Show the oracle version converges to* 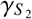. The *w*-weighted sample in *S*_1_ approximates the *S*_2_ covariate distribution, because by Bayes’ theorem:

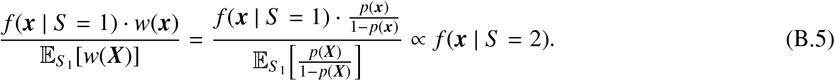

This is the standard odds-ratio reweighting identity [8]. Therefore, for any measurable function *h*(***X***) bounded by (R1) and (R3):

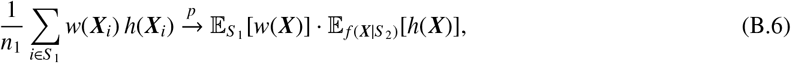

by the law of large numbers applied to i.i.d. draws from *S*_1_.

Applying this to the numerator and denominator of 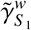, and using Assumption 3 and 6 to write:

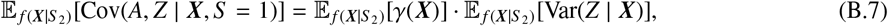

the ratio telescopes to:

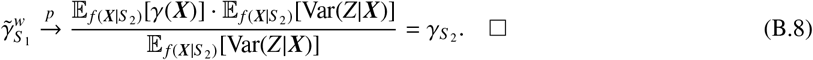

*Key role of assumptions*.Assumption 6 (Conditional Transportability) is used in Step 2 to equate *γ*(***x***) in *S*_1_ with that in *S*_2_ conditional on ***X*** = ***x***. Assumption 7 guarantees the odds-ratio reweighting identity is well-defined. If either fails, 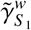 need not converge to 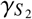.

#### Lemma 3 (Consistency of Reduced-Form Estimator).

*Under Assumptions 1, 2, and 4, and (R3):*

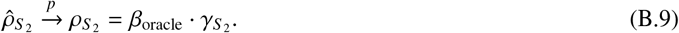

#### Proof.

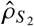 is the OLS coefficient on *Z* in the regression of *Y* on (1, *Z*, ***X***) in *S*_2_. By (R3) and standard OLS consistency [12], 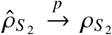, where 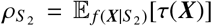 by the exclusion restriction (Assumption 2). The factorisation 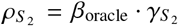 follows from the definition of *β*_oracle_ in equation (6) and the definition of *τ*(***x***) = *λ*(***x***) · *γ*(***x***).

#### Proof of Proposition 1.

By Lemma 2, 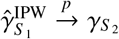. By Lemma 3, 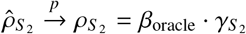. By (R4), 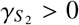, so the ratio is well-defined in a neighborhood of the probability limit. The continuous mapping theorem then gives:

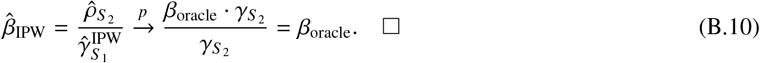

## Appendix C. Data-Generating Process Details

### Appendix C.1. Covariate Generation (Super-Population)

State attributes for *K* = 50 states are generated as follows:

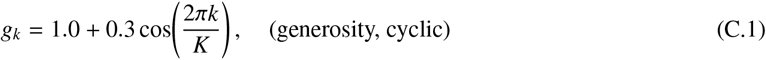

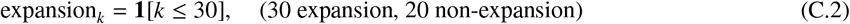

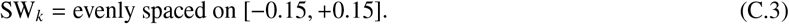

State probabilities follow a cosine-modulated distribution: Pr[state = *k*] ∝ 3 + cos(2*πk*/*K*) for *k* = 1, …, 50.

Full covariate generation follows the pseudocode in Section 7. The truncated normal for age uses rejection sampling. Education is drawn via a sequential probit parameterization to maintain valid probability simplex.

### Appendix C.2. Default Parameters

**Table C.9.** Default DGP parameter values.

| Parameter | Symbol | Value | Role |
| --- | --- | --- | --- |
| True LATE | $\beta_{\text{oracle}}$ | 2.0 | Causal estimand |
| First-stage intercept | $\pi_0$ | 0.60 | Baseline compliance |
| Income slope in $\gamma(X)$ | $\pi_{\text{inc}}$ | -0.03 | Primary heterogeneity driver |
| Education slope | $\pi_{\text{edu}}$ | 0.10 | Awareness effect |
| Family size slope | $\pi_{\text{fam}}$ | 0.15 | Household compliance effect |
| Age slope | $\pi_{\text{age}}$ | -0.004 | Stability effect |
| State generosity slope | $\pi_{\text{state}}$ | 0.05 | Policy environment |
| Floor on $\gamma(X)$ | $\delta_{\text{min}}$ | 0.05 | Ensures all compliers |
| S2 intercept | $\alpha_0$ | 1.5 | Baseline Medicaid enrollment |
| S2 income selection | $\alpha_{\text{inc}}$ | -2.5 | Medicaid income threshold |
| S2 expansion states | $\alpha_{\text{state}}$ | 0.8 | Expansion enrollment |
| S2 sex | $\alpha_{\text{sex}}$ | 0.3 | Female enrollment differential |
| S2 race (Black) | $\alpha_{\text{rB}}$ | 0.2 | Racial enrollment differential |
| S2 race (Hispanic) | $\alpha_{\text{rH}}$ | 0.15 | Racial enrollment differential |
| S2 family size | $\alpha_{\text{fam}}$ | 0.4 | Family coverage effect |
| S2 age | $\alpha_{\text{age}}$ | -0.01 | Age enrollment pattern |
| S1 intercept | $\beta_0$ | 0.0 | Survey baseline |
| S1 income selection | $\beta_{\text{inc}}$ | -0.3 | Mild low-income oversample |
| S1 age (< 30) | $\beta_{\text{age}}$ | 0.2 | Young adult oversample |
| S1 race (Hispanic) | $\beta_{\text{rH}}$ | 0.15 | ACS Hispanic oversample |
| Outcome effect | $\beta_{\text{oracle}}$ | 2.0 | Same as LATE |
| Direct income $\rightarrow Y$ | $\delta_{\text{inc}}$ | -0.5 | Confounding |
| Direct education $\rightarrow Y$ | $\delta_{\text{edu}}$ | -0.3 | Confounding |
| Direct age $\rightarrow Y$ | $\delta_{\text{age}}$ | 0.04 | Confounding |
| Direct state $\rightarrow Y$ | $\delta_{\text{state}}$ | 0.2 | Geographic confounding |
| Treatment noise | $\sigma_A$ | 1.5 | First-stage residual |
| Outcome noise | $\sigma_Y$ | 3.0 | Outcome residual |
| Super-population | $N$ | 500,000 | Monte Carlo precision |
| S1 sample size | $n_1$ | 3,000 | Survey |
| S2 sample size | $n_2$ | 100,000 | Claims |
| States | $K$ | 50 | Cluster count |
| Simulations | $N_{\text{sim}}$ | 500 | Monte Carlo precision |
| Bootstrap replicates | $B$ | 200 | Bootstrap precision |

## Appendix D. Computational Implementation Notes

The simulation study uses Python 3.12 with NumPy, SciPy, and pandas, employing custom cluster-robust sandwich estimators for computational efficiency. The empirical application uses the same Python environment. Key implementation choices:

### Custom cluster-robust sandwich estimators

We implement `fast_cluster_ols` and `fast_cluster_wls` with hand-rolled HC1-cluster sandwich matrices. This avoids the overhead of generic robust regression packages in the inner Monte Carlo loop, reducing runtime by approximately 40%.

### IPW weights

The IPW correction in the empirical application estimates propensity scores via logistic regression of sample membership *S* ∈ {1, 2} on pooled covariates, with weights 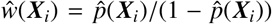 for *i* ∈ *S*_1_ normalized to mean unity after 99th-percentile trimming. The logistic model is fitted via iteratively reweighted least squares. Bootstrap replicates re-estimate the propensity model from scratch in each replicate to propagate weight-estimation uncertainty.

### Parallelization

Each simulation scenario runs *N*_sim_ replicates in parallel on 10 cores using Python’s `multiprocessing.Pool` with a fork-based initializer pattern to avoid serializing the super-population array. Each replicate uses a deterministic seed for reproducibility.

### Replication

All code is implemented in Python 3.12 and is available upon request.

## References

[1] Joshua D Angrist and Alan B Krueger. The effect of age at school entry on educational attainment: an application of instrumental variables with moments from two samples. Journal of the American statistical Association, 87 (418):328–336, 1992.

[2] Joshua D Angrist and Alan B Krueger. Split-sample instrumental variables estimates of the return to schooling. Journal of Business & Economic Statistics, 13(2):225–235, 1995.

[3] Elias Bareinboim and Judea Pearl. Causal inference and the data-fusion problem. Proceedings of the National Academy of Sciences, 113(27):7345–7352, 2016.

[4] Bénédicte Colnet, Imke Mayer, Guanhua Chen, Awa Dieng, Ruohong Li, Gaël Varoquaux, Jean-Philippe Vert, Julie Josse, and Shu Yang. Causal inference methods for combining randomized trials and observational studies: a review. Statistical science: a review journal of the Institute of Mathematical Statistics, 39(1):165, 2024.

[5] Issa J Dahabreh, Jon A Steingrimsson, James M Robins, and Miguel A Hernán. Randomized trials and their observational emulations: a framework for benchmarking and joint analysis. arXiv preprint arXiv:2203.14857, 2022.

[6] Thomas S Dee and William N Evans. Teen drinking and educational attainment: evidence from two-sample instrumental variables estimates. Journal of Labor Economics, 21(1):178–209, 2003.

[7] Amy Finkelstein and Katherine Baicker. Oregon health insurance experiment, 2007–2010, 2014. URL https://www.icpsr.umich.edu/web/HMCA/studies/34314.

[8] Keisuke Hirano, Guido W Imbens, and Geert Ridder. Efficient estimation of average treatment effects using the estimated propensity score. Econometrica, 71(4):1161–1189, 2003.

[9] Atsushi Inoue and Gary Solon. Two-sample instrumental variables estimators. The Review of Economics and Statistics, 92(3):557–561, 2010.

[10] Sarah Miller, Norman Johnson, and Laura R Wherry. Medicaid and mortality: new evidence from linked survey and administrative data. The Quarterly Journal of Economics, 136(3):1783–1829, 2021.

[11] Whitney K. Newey. The asymptotic variance of semiparametric estimators. Econometrica, 62(6):1349–1382, 1994.

[12] Whitney K. Newey and Daniel McFadden. Large sample estimation and hypothesis testing. In Robert F. Engle and Daniel L. McFadden, editors, Handbook of Econometrics, volume 4, pages 2111–2245. Elsevier, 1994.

[13] David Pacini and Frank Windmeijer. Robust inference for the two-sample 2sls estimator. Economics letters, 146:50–54, 2016.

[14] Judea Pearl and Elias Bareinboim. Transportability of causal and statistical relations: A formal approach. In Proceedings of the AAAI Conference on Artificial Intelligence, volume 25, pages 247–254, 2011.

[15] Xu Shi, Ziyang Pan, and Wang Miao. Data integration in causal inference. Wiley Interdisciplinary Reviews: Computational Statistics, 15(1):e1581, 2023.

[16] Sarah L Taubman, Heidi L Allen, Bill J Wright, Katherine Baicker, and Amy N Finkelstein. Medicaid increases emergency-department use: evidence from oregon’s health insurance experiment. Science, 343(6168):263–268, 2014.

[17] Qingyuan Zhao, Jingshu Wang, Wes Spiller, Jack Bowden, and Dylan S Small. Two-sample instrumental variable analyses using heterogeneous samples. Statistical Science, 34(2):317–333, 2019.

